# The effectiveness of the ‘RecoverEsupport’ digital health intervention in enhancing recovery from colorectal cancer surgery: RCT outcomes from the hospital admission

**DOI:** 10.64898/2026.11.22.26368447

**Authors:** Rebecca Wyse, Priscilla Viana da Silva, Anna Palazzi-Parsons, Emma Sansalone, Erin Forbes, Sancha Robinson, Sally-Ann Johnston, Kristy Fakes, Erin Nolan, Lucy Leigh, Julie Charlton, Steven Smith

## Abstract

**Background:** Enhanced Recovery After Surgery (ERAS) guidelines improve recovery outcomes following colorectal (CR) cancer surgery. However, adherence to patient-led ERAS behaviours within the guidelines remains variable. Digital health interventions (DHIs) may increase adherence to these behaviours during the perioperative period. This study evaluated the effectiveness of a behaviourally-informed DHI – RecoverEsupport – in increasing adherence and improving recovery outcomes following colorectal cancer surgery.

**Methods:** A parallel-group RCT was conducted in two Australian hospitals. Adults undergoing colorectal cancer surgery were randomised (1:1) to usual care or usual care plus the RecoverEsupport DHI targeting ERAS behaviours, including early mobilisation and early oral fluid and food intake. Between-group differences in the primary (length of hospital stay (LOS)) and secondary outcomes (patient-led ERAS behaviours and quality of recovery (QoR-15)) were investigated using regression analyses within a Bayesian framework.

**Results:** On Day 1 post-surgery, there was moderate to strong evidence that the intervention was superior in increasing time out of bed [median 195 vs 60 minutes; RR=2.38, 95% CrI 1.43–3.29; Pr(Superiority)=1.000], and fluid intake [75% vs 45% consuming ≥4 cups; OR=5.67, 95% CrI 1.12–30.1; Pr(Superiority)=0.987], and moderate evidence of a small increase in time spent moving around [median 23 vs 20 minutes; RR=1.28, 95% CrI 0.9-1.86; Pr(Superiority)=0.904]. There was also moderate evidence of intervention superiority in terms of quality of recovery [mean QoR-15 difference=11.64, 95% CrI −2.97–25.63; Pr(Superiority)=0.937] exceeding the clinically meaningful difference.

**Conclusions:** Although the RecoverEsupport intervention did not reduce length of hospital stay, it increased early mobilisation, fluid intake and quality of recovery for CR surgery patients.

## INTRODUCTION

Each year, approximately 2 million people worldwide are diagnosed with colorectal (CR) cancer, with most patients undergoing surgery.^1,2^ Enhanced Recovery After Surgery (ERAS) guidelines provide evidence-based recommendations to optimise recovery following CR surgery.^3^ Systematic reviews demonstrate that implementing ERAS guidelines is associated with reduced length of hospital stay and complications.^4,5^ ERAS guidelines comprise both clinician- and patient-led behaviours. ERAS guidelines include preoperative (e.g. alcohol and smoking cessation/reduction) and postoperative behaviours (e.g. early mobilisation, oral food and fluid intake).^6^

Adherence to patient-led ERAS behaviours remains inconsistent.^7^ Although patients are not solely responsible for these behaviours, they typically have greater control and agency over them compared to other ERAS guidelines, making them particularly amenable to patient-facing behavioural interventions. Digital health interventions (DHIs) may enable CR patients to directly receive such support throughout the perioperative period. Emerging evidence suggests DHIs in this context are feasible to deliver, acceptable to patients,^8,9^ and can improve early mobilisation^10,11^ and oral intake^11^ but, there is only limited evidence (from a non-randomised trial) that such interventions improve length of stay.^12^ The few randomised trials conducted to-date have been limited either by interventions targeting only a narrow range of ERAS behaviours^10^ or by a failure to ground the intervention in behavioural science.^11,13^ RecoverEsupport, a behaviourally-informed DHI targeting a suite of patient-led ERAS behaviours was developed to address these limitations. It was hypothesised that the RecoverEsupport DHI would improve clinical, behavioural and patient-reported outcomes relative to usual care.

### AIM

The study aim was to evaluate the effectiveness of RecoverEsupport in supporting recovery following colorectal cancer surgery, as assessed via hospital length of stay (primary outcome), patient adherence to ERAS behaviours and quality of recovery (secondary outcomes).

## METHODS

### Design

This was a parallel-group, superiority, randomised controlled trial. All participants received usual care, and half were randomised to also receive the RecoverEsupport DHI. The trial was prospectively registered (ACTRN12621001533886; 10/11/2021) with methods fully described in the trial protocol^14^ and outlined below, and is reported according to the CONSORT checklist (Supplementary File 1). This paper reports on the outcomes collected during participants’ hospital admission following their surgery.

### Ethics and Governance

Approval was provided by the Human Research Ethics Committees (HREC) of the Hunter New England Local Health District (2019/ETH00869), the University of Newcastle (H-2015-0364) and the hospital sites.

### Setting

Participants were adult patients undergoing colorectal surgery at one of two hospitals within Newcastle, NSW, Australia. Hospital 1 was a large public tertiary hospital, and Hospital 2 was a private hospital co-located on the public hospital grounds.

### Eligibility criteria

Eligible patients: were aged ≥18 years; had a planned surgery for colorectal cancer; with an expected inpatient stay of at least 3 days; had Internet access; were free from cognitive impairment; were physically and emotionally capable; and were literate in English.

### Exclusion Criteria

Patients with advanced disease (metastasis) and taking daily opioids were excluded. Patients identified within 3 days of surgery were not invited into the trial, as there was insufficient time to expose them to the intervention strategies prior to surgery.

### Recruitment

Potentially eligible patients were identified from the surgical lists at each hospital. Hospital staff screened and invited eligible patients to participate either by telephone or face-to-face at the preoperative clinic or presurgical appointment. Participants consented by completing the hard-copy or online consent form. Recruitment and data collection commenced at the public hospital in July 2022 and at the private hospital in August 2023 and continued until October 2025.

### Randomisation

Participants were randomised by an independent data scientist via REDCap in a 1:1 ratio to the intervention or control group, using a block randomisation schedule with block size varying between 4-6. Randomisation occurred as soon as the baseline survey was completed, or 3 days prior to surgery, whichever occurred first. Participants were automatically allocated to their group within REDCap. Due to the nature of the behavioural intervention, patients and nursing staff were not blinded to group allocation.

### Usual Care

All CR surgery patients receive preoperative information and support through clinics and/or preoperative appointments (either face-to-face or via telephone) during which time they may see their surgeon, anaesthetist and/or specialty nurses.

### The RecoverEsupport Intervention

#### Overview

Patients randomised to the treatment group received access to the RecoverEsupport program, a bespoke, behaviourally-informed DHI designed to support adherence to patient-led ERAS behaviours for colorectal surgery. The intervention consisted of a website and a series of reminders delivered via REDCap (Figure 1). It predominantly targeted postoperative ERAS behaviours using evidence-based behaviour change strategies (Supplementary File 2). RecoverEsupport was developed using co-design principles with behavioural scientists, clinicians and patients. For more details, see the published trial protocol.^14^

**FIGURE 1.**
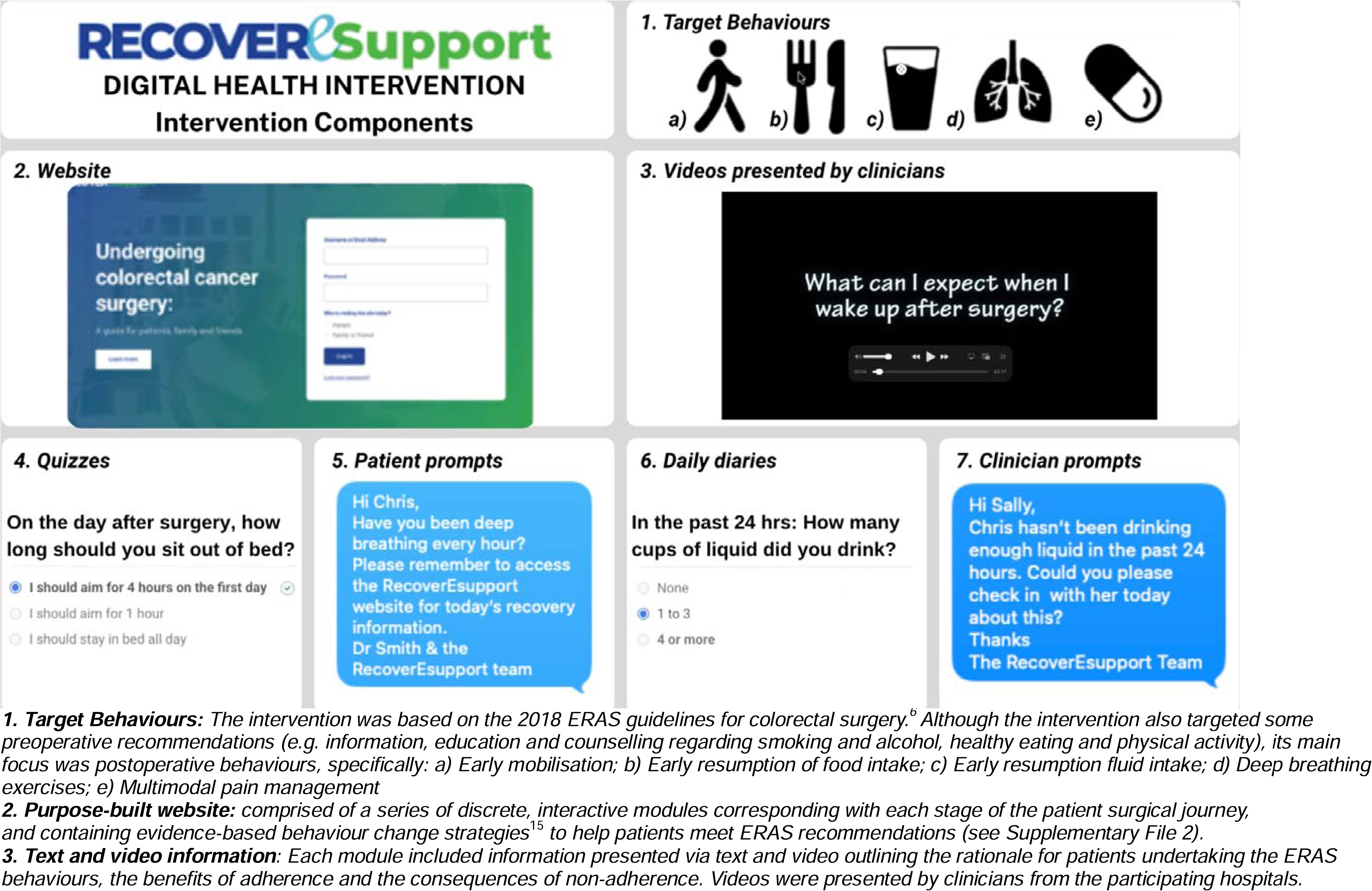
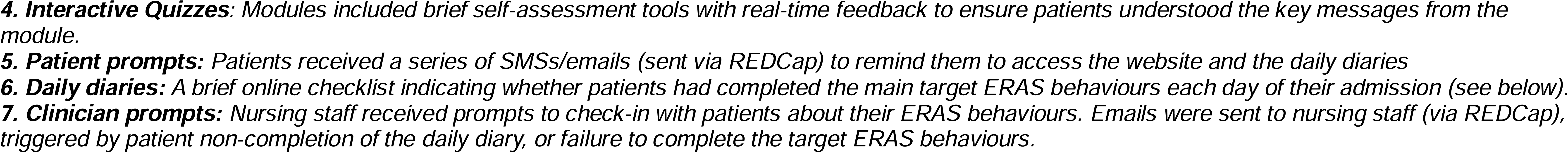
Target behaviours and main components of the RecoverEsupport Digital Health Intervention to enhance surgical recovery in colorectal cancer surgery patients.

**FIGURE 2.**
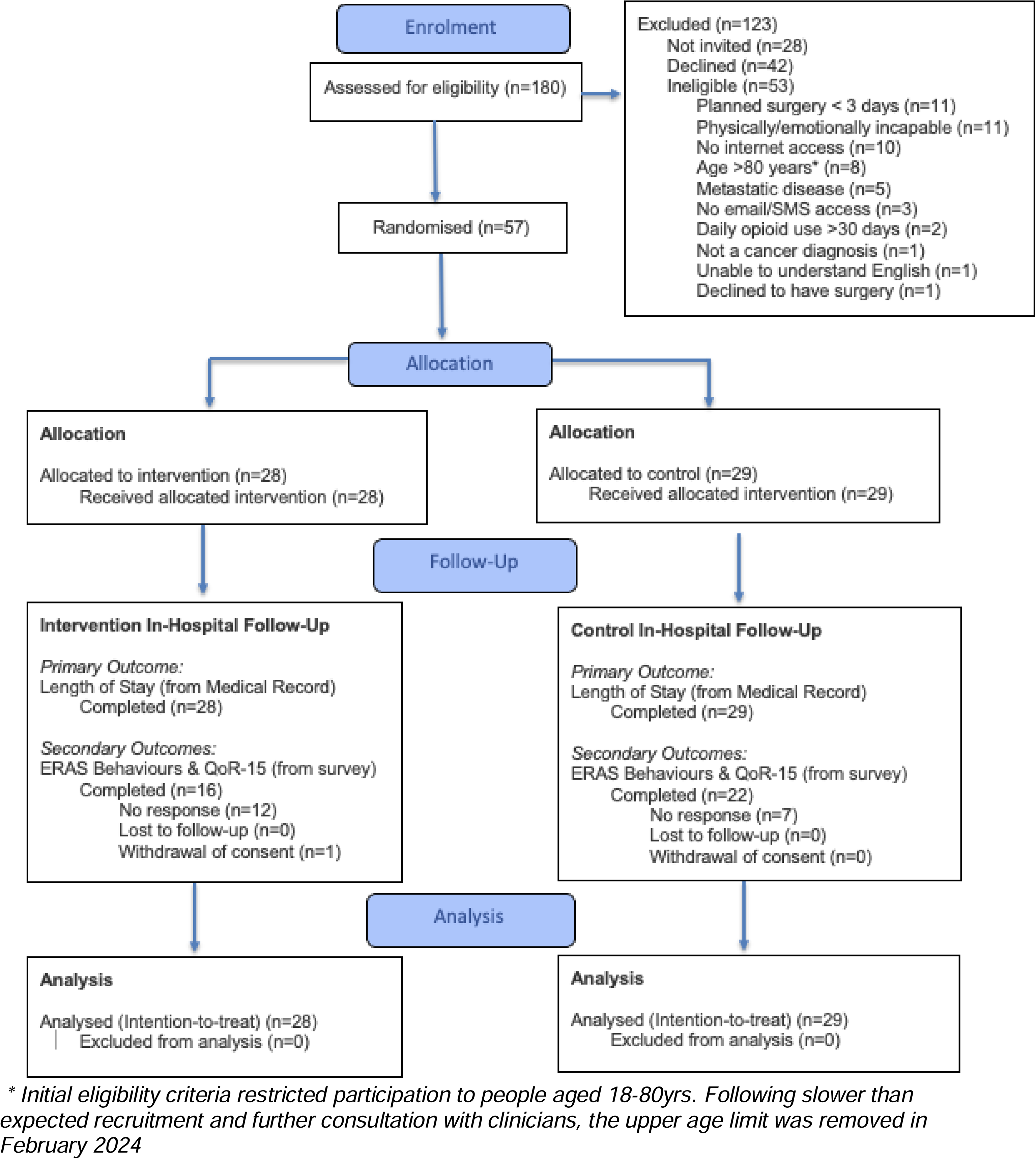
CONSORT flowchart showing participant progression through the RecoverEsupport RCT.

### Data Collection

Data was collected from patient medical records and online patient surveys and stored securely in a REDCap database. Patients were invited to complete online surveys at baseline and in hospital (Day 2 post-surgery) and if these were not completed, patients were sent email/SMS reminders according to a pre-specified schedule.

### Outcomes

#### Primary outcome

The primary trial outcome was the length of hospital stay in days (i.e. the difference between discharge and admission date), based on the medical record.

#### Secondary outcomes

Patient-reported outcomes were assessed via the online survey administered on Day 2. (Additional outcomes were collected 1- to 3-months post-discharge and are reported separately.)

##### Patient ERAS behaviours and knowledge

Items were specifically developed for this trial to assess adherence to the ERAS patient-led behaviours and ERAS knowledge. The survey asked patients to report on their first full day on the ward after surgery (henceforth referred to as ‘Day 1’ behaviours etc). See Supplementary file 3 for the full wording of items.

##### Quality of Recovery

The QoR-15 tool was used to assess the quality of patient recovery in the previous 24 hours whilst in hospital (i.e. Day 1). This 15-item patient-reported tool is responsive, valid, and reliable.^16^

### Patient demographics and treatment characteristics

Patient characteristics were extracted from the patient medical record by hospital staff and patient information collected from the baseline survey. Patients recruited within 5 days of their surgery date were not invited to complete the baseline survey and their characteristics were taken exclusively from the medical record.

#### Intervention Fidelity

Summary statistics on intervention engagement were calculated from paradata from the RecoverEsupport website and REDCap.

#### Sample size

The sample size was based on a mean length of stay of 8.6 days (SD=7.6) assuming 75% eligibility, 85% consent rate, and 10% attrition (unpublished pilot and medical record data from Hospital 1). 167 participants per arm would provide 80% power to detect a between-group decrease in mean length of stay of 2.5 days, which is similar to the decrease observed in a previous (non-randomised) trial.^17^

#### Analysis

Descriptive statistics were calculated using means (SD) and percentages as appropriate. The target sample size was not achieved and the analysis was conducted within a Bayesian Framework^18^ given it is flexible and robust, and allows probabilistic statements about the direction and magnitude of effects (i.e. provides a probability of the intervention being superior to the control), supporting better decision-making in smaller trials. This contrasts with the frequentist framework, where inference of intervention superiority is often reduced to a binary judgement of statistical significance (i.e. superior / not superior, p-value < 0.05).

An Intention-to-Treat (ITT) approach was used for the primary outcome (LOS) and secondary outcomes (ERAS behaviours and QoR). Multiple imputation was used to impute missing secondary outcome data under a missing at random (MAR) assumption using the ‘mice’ package.^19^ A per protocol (PP) analysis and sub-group analyses were also conducted for the primary outcome. Descriptive statistics were calculated for LOS stratified by sub-groups of age (based on a median split), gender (female; male), and hospital type (public; private) (Supplementary File 4).

Between-group differences across all outcomes were assessed independently within a Bayesian framework using the brms package.^20–23^ All regression models included group allocation (intervention/control), hospital site (public/private), presence of stoma (yes/no), and surgery type (open/laparoscopic) as fixed effects. Intervention effect estimates were reported with 95% credible intervals based on the highest posterior density and the one-sided probability that the intervention was superior (Pr(Superiority)). Under a Bayesian approach, posterior probabilities do not have widely agreed-upon interpretive cut-points.^24^ However, consistent with previously published Bayesian analysis, probabilities of superiority exceeding 0.90 and 0.95 can be viewed as moderate and stronger support for intervention superiority respectively^25^ but should not be considered dichotomous cutoffs as per a frequentist approach. The statistician was blinded to group allocation.

#### Protocol amendments

In order to increase the number of participants, the requirement that the baseline survey be completed was dropped, given the primary outcome and key demographic and adjustment variables were all available from the medical record. Minor amendments to the analysis in the published protocol were required. Analysis by cancer stage was initially planned, but given this information was not consistently available for patients, this was not conducted. Due to slower than expected recruitment, a second hospital was added in August 2023. The analysis was adjusted to incorporate hospital site. However, in order to minimise model overfitting (and following an assessment of the effective number of parameters in the ‘loo’ package)^26^ baseline QoL was excluded from the analysis. Due to the lower than expected sample size, instead of the proposed subgroup analysis, we descriptively present the median LOS by age (based on a median split), gender (identification as female vs male), and hospital type (public vs private). (Supplementary File 4).

## RESULTS

In total, 180 patients were screened, with 28 not approached in clinic (missed), 53 not meeting eligibility criteria, and 57 out of 99 eligible patients consenting (58%). Recruitment was stopped based on the funding timeline, despite not having reached the target. Consenters and non-consenters were similar in terms of age and gender. 27 of 52 (52%) and 30 of 47 (64%) invited and eligible patients were recruited from the public and private hospital respectively. Figure 3 shows the flow of participants through the trial.

Participants’ baseline demographics and medical characteristics are reported in Table 1. Despite the small sample size, baseline characteristics appeared reasonably balanced between groups with the exception of ASA, which indicate a higher proportion of less healthy participants in the intervention group relative to the control group (75% vs 46% with ASA III; severe systematic disease [no statistical testing conducted]).

**TABLE 11:**
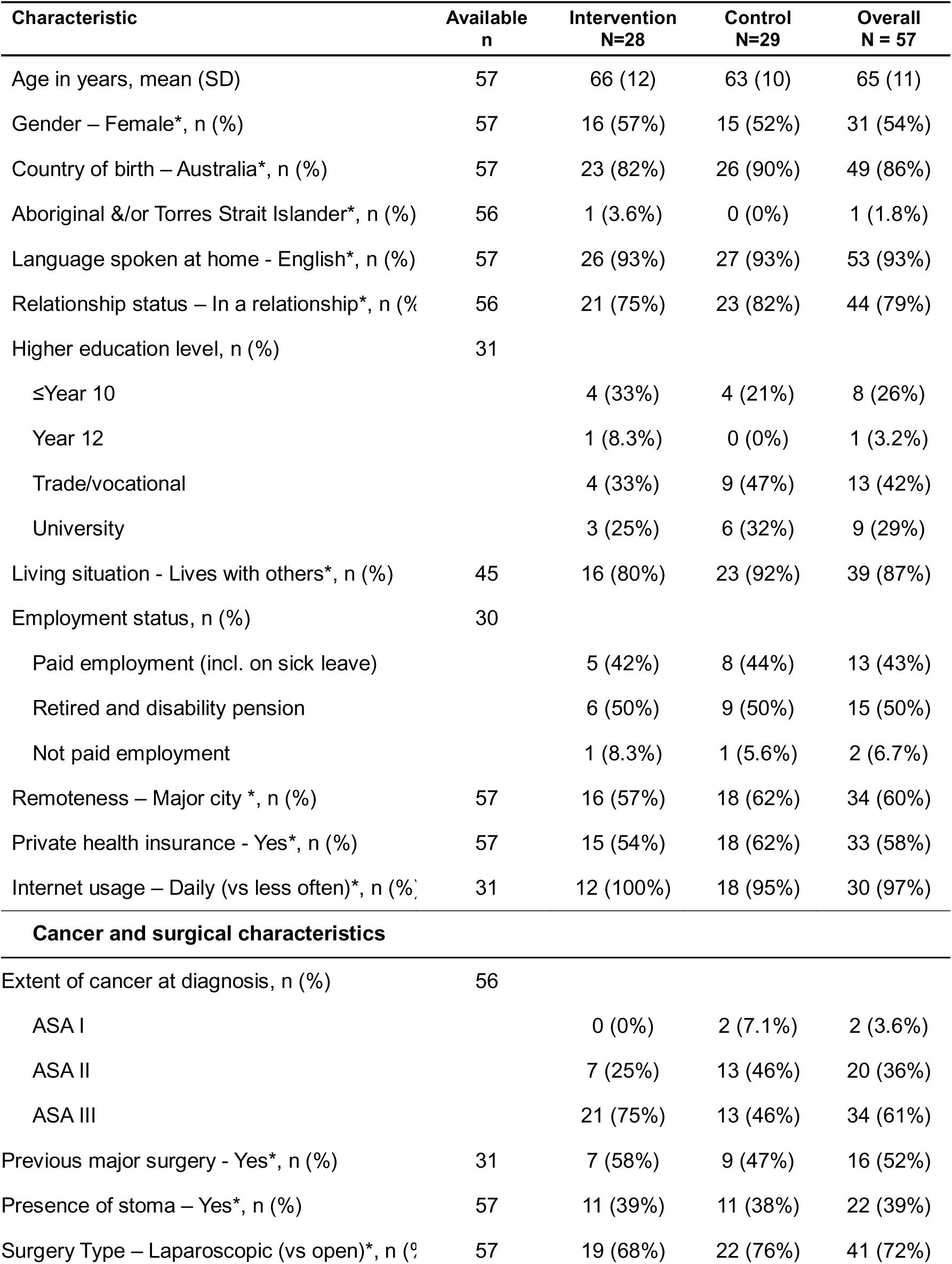
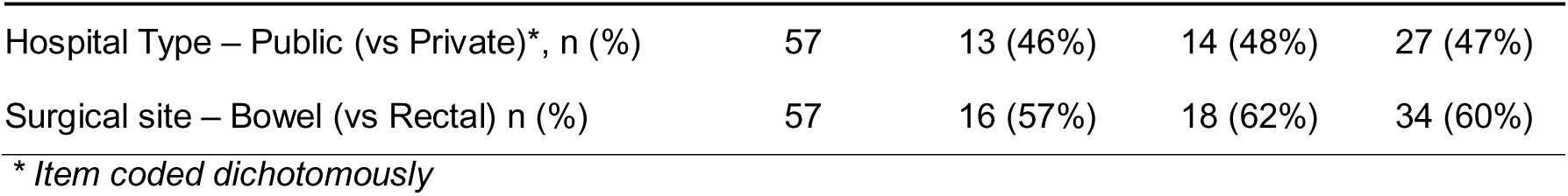
Characteristics of intervention and control participants and the overall sample in the RecoverEsupport RCT.

| Characteristic | Available<br>n | Intervention<br>N=28 | Control<br>N=29 | Overall<br>N = 57 |
|---|---|---|---|---|
| Age in years, mean (SD) | 57 | 66 (12) | 63 (10) | 65 (11) |
| Gender – Female*, n (%) | 57 | 16 (57%) | 15 (52%) | 31 (54%) |
| Country of birth – Australia*, n (%) | 57 | 23 (82%) | 26 (90%) | 49 (86%) |
| Aboriginal &/or Torres Strait Islander*, n (%) | 56 | 1 (3.6%) | 0 (0%) | 1 (1.8%) |
| Language spoken at home - English*, n (%) | 57 | 26 (93%) | 27 (93%) | 53 (93%) |
| Relationship status – In a relationship*, n (%[illegible] | 56 | 21 (75%) | 23 (82%) | 44 (79%) |
| Higher education level, n (%) | 31 |  |  |  |
| ≤Year 10 |  | 4 (33%) | 4 (21%) | 8 (26%) |
| Year 12 |  | 1 (8.3%) | 0 (0%) | 1 (3.2%) |
| Trade/vocational |  | 4 (33%) | 9 (47%) | 13 (42%) |
| University |  | 3 (25%) | 6 (32%) | 9 (29%) |
| Living situation - Lives with others*, n (%) | 45 | 16 (80%) | 23 (92%) | 39 (87%) |
| Employment status, n (%) | 30 |  |  |  |
| Paid employment (incl. on sick leave) |  | 5 (42%) | 8 (44%) | 13 (43%) |
| Retired and disability pension |  | 6 (50%) | 9 (50%) | 15 (50%) |
| Not paid employment |  | 1 (8.3%) | 1 (5.6%) | 2 (6.7%) |
| Remoteness – Major city *, n (%) | 57 | 16 (57%) | 18 (62%) | 34 (60%) |
| Private health insurance - Yes*, n (%) | 57 | 15 (54%) | 18 (62%) | 33 (58%) |
| Internet usage – Daily (vs less often)*, n (%) | 31 | 12 (100%) | 18 (95%) | 30 (97%) |
| **Cancer and surgical characteristics** |  |  |  |  |
| Extent of cancer at diagnosis, n (%) | 56 |  |  |  |
| ASA I |  | 0 (0%) | 2 (7.1%) | 2 (3.6%) |
| ASA II |  | 7 (25%) | 13 (46%) | 20 (36%) |
| ASA III |  | 21 (75%) | 13 (46%) | 34 (61%) |
| Previous major surgery - Yes*, n (%) | 31 | 7 (58%) | 9 (47%) | 16 (52%) |
| Presence of stoma – Yes*, n (%) | 57 | 11 (39%) | 11 (38%) | 22 (39%) |
| Surgery Type – Laparoscopic (vs open)*, n ([illegible] | 57 | 19 (68%) | 22 (76%) | 41 (72%) |
| Hospital Type – Public (vs Private)*, n (%) | 57 | 13 (46%) | 14 (48%) | 27 (47%) |
| Surgical site – Bowel (vs Rectal) n (%) | 57 | 16 (57%) | 18 (62%) | 34 (60%) |

*\* Item coded dichotomously*

### Length of Stay

Primary outcome data was available for all participants. Given the skewed data, the primary outcome analysis used medians instead of means. The crude median LOS in each group was 6 days (IQR [4,9] and [4,8] for intervention and control group respectively) (range: 3 to 62 days). There was a low probability of intervention superiority relative to controls, with the point estimate indicating 7% greater length of stay for the intervention compared to the control (RR=1.07; 95% CrI 0.9-1.28; Pr(Superiority)=0.223) (Table 2). For descriptive purposes, the overall mean LOS was 8.8 days (SD=9.4); 9.4 days (SD=11.3) and 8.1 days (SD=7.0) for the intervention and control group respectively. See Supplementary File 4 for median LOS by gender, age and hospital type.

**TABLE 2:**
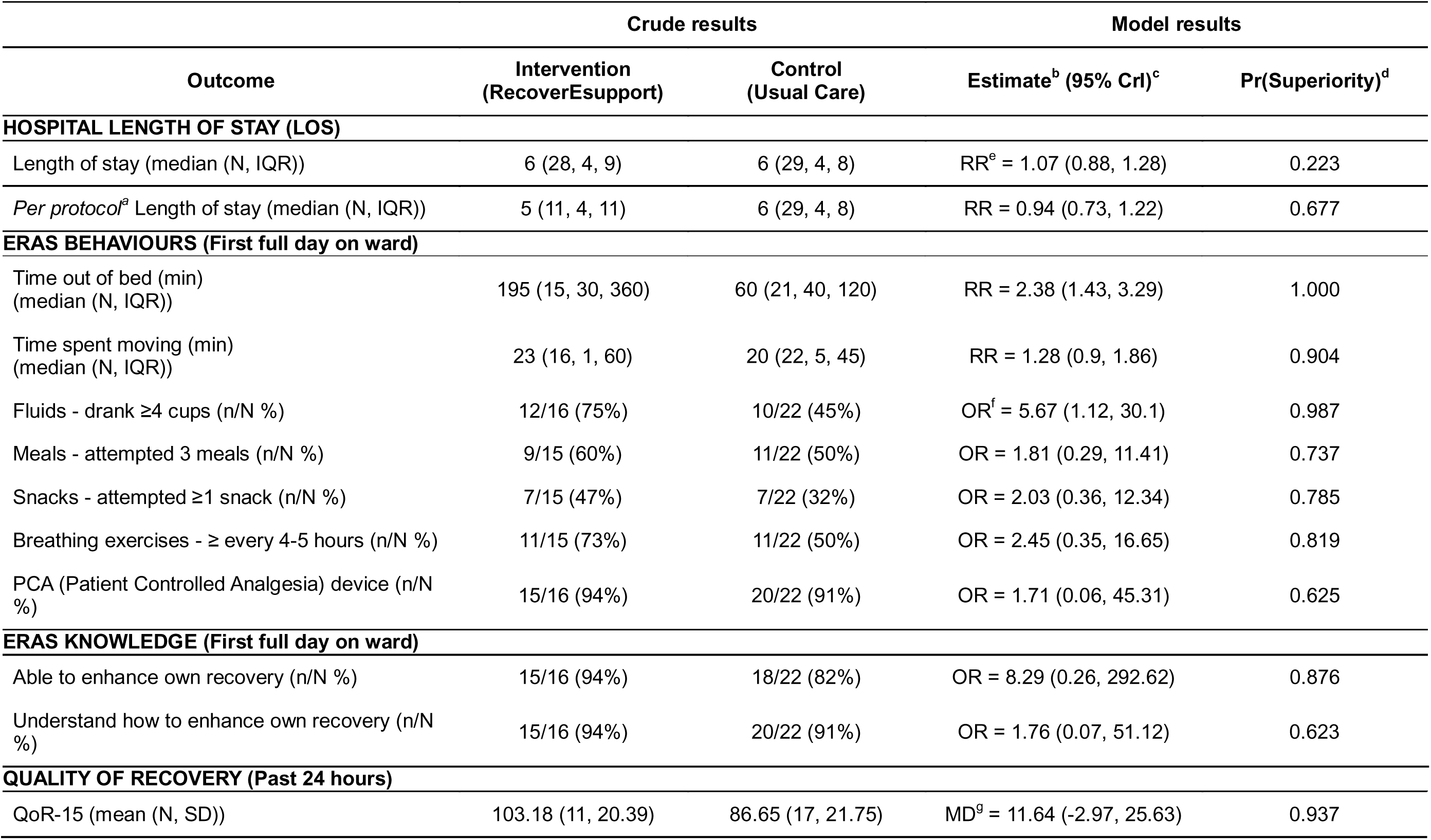
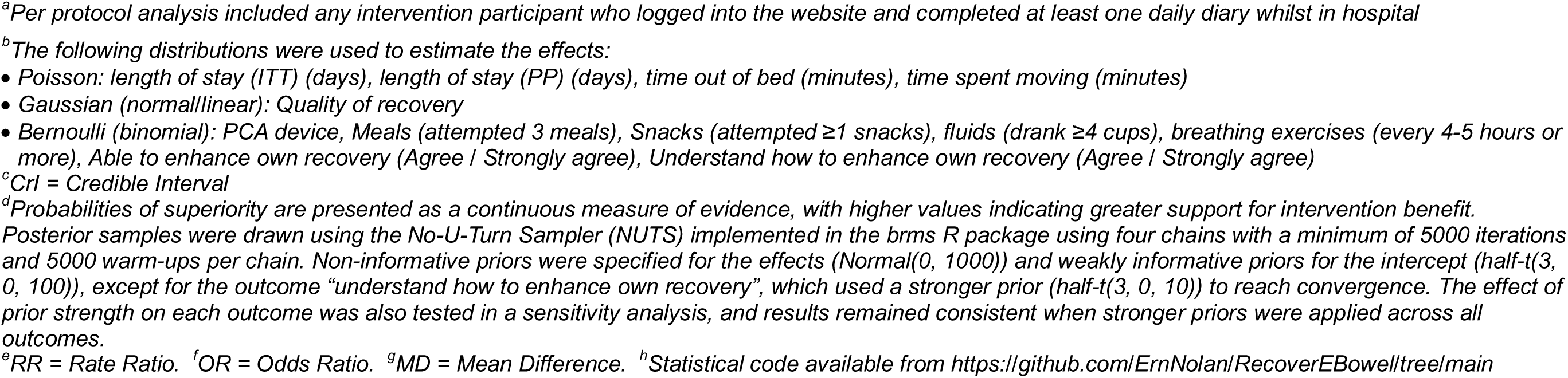
Summary of crude and model results for all ITT outcomes in the RecoverEsupport RCT^h^.

| Outcome | Crude results |  | Model results |  |
|---|---|---|---|---|
| Outcome | Intervention<br>(RecoverEsupport) | Control<br>(Usual Care) | Estimate<sup>b</sup> (95% CrI)<sup>c</sup> | Pr(Superiority)<sup>d</sup> |
| **HOSPITAL LENGTH OF STAY (LOS)** |  |  |  |  |
| Length of stay (median (N, IQR)) | 6 (28, 4, 9) | 6 (29, 4, 8) | RR<sup>e</sup> = 1.07 (0.88, 1.28) | 0.223 |
| *Per protocol*<sup>a</sup> Length of stay (median (N, IQR)) | 5 (11, 4, 11) | 6 (29, 4, 8) | RR = 0.94 (0.73, 1.22) | 0.677 |
| **ERAS BEHAVIOURS (First full day on ward)** |  |  |  |  |
| Time out of bed (min)<br>(median (N, IQR)) | 195 (15, 30, 360) | 60 (21, 40, 120) | RR = 2.38 (1.43, 3.29) | 1.000 |
| Time spent moving (min)<br>(median (N, IQR)) | 23 (16, 1, 60) | 20 (22, 5, 45) | RR = 1.28 (0.9, 1.86) | 0.904 |
| Fluids - drank ≥4 cups (n/N %) | 12/16 (75%) | 10/22 (45%) | OR<sup>f</sup> = 5.67 (1.12, 30.1) | 0.987 |
| Meals - attempted 3 meals (n/N %) | 9/15 (60%) | 11/22 (50%) | OR = 1.81 (0.29, 11.41) | 0.737 |
| Snacks - attempted ≥1 snack (n/N %) | 7/15 (47%) | 7/22 (32%) | OR = 2.03 (0.36, 12.34) | 0.785 |
| Breathing exercises - ≥ every 4-5 hours (n/N %) | 11/15 (73%) | 11/22 (50%) | OR = 2.45 (0.35, 16.65) | 0.819 |
| PCA (Patient Controlled Analgesia) device (n/N %) | 15/16 (94%) | 20/22 (91%) | OR = 1.71 (0.06, 45.31) | 0.625 |
| **ERAS KNOWLEDGE (First full day on ward)** |  |  |  |  |
| Able to enhance own recovery (n/N %) | 15/16 (94%) | 18/22 (82%) | OR = 8.29 (0.26, 292.62) | 0.876 |
| Understand how to enhance own recovery (n/N %) | 15/16 (94%) | 20/22 (91%) | OR = 1.76 (0.07, 51.12) | 0.623 |
| **QUALITY OF RECOVERY (Past 24 hours)** |  |  |  |  |
| QoR-15 (mean (N, SD)) | 103.18 (11, 20.39) | 86.65 (17, 21.75) | MD<sup>g</sup> = 11.64 (-2.97, 25.63) | 0.937 |

<sup>a</sup>*Per protocol analysis included any intervention participant who logged into the website and completed at least one daily diary whilst in hospital*
<sup>b</sup>*The following distributions were used to estimate the effects:*
• *Poisson: length of stay (ITT) (days), length of stay (PP) (days), time out of bed (minutes), time spent moving (minutes)*
• *Gaussian (normal/linear): Quality of recovery*
• *Bernoulli (binomial): PCA device, Meals (attempted 3 meals), Snacks (attempted ≥1 snacks), fluids (drank ≥4 cups), breathing exercises (every 4-5 hours or more), Able to enhance own recovery (Agree / Strongly agree), Understand how to enhance own recovery (Agree / Strongly agree)*
<sup>c</sup>*CrI = Credible Interval*
<sup>d</sup>*Probabilities of superiority are presented as a continuous measure of evidence, with higher values indicating greater support for intervention benefit.*
*Posterior samples were drawn using the No-U-Turn Sampler (NUTS) implemented in the brms R package using four chains with a minimum of 5000 iterations and 5000 warm-ups per chain. Non-informative priors were specified for the effects (Normal(0, 1000)) and weakly informative priors for the intercept (half-t(3, 0, 100)), except for the outcome “understand how to enhance own recovery”, which used a stronger prior (half-t(3, 0, 10)) to reach convergence. The effect of prior strength on each outcome was also tested in a sensitivity analysis, and results remained consistent when stronger priors were applied across all outcomes.*
<sup>e</sup>*RR = Rate Ratio.* <sup>f</sup>*OR = Odds Ratio.* <sup>g</sup>*MD = Mean Difference.* <sup>h</sup>*Statistical code available from https://github.com/ErnNolan/RecoverEBowell/tree/main*

#### Per protocol analysis

Eleven intervention participants (39%) engaged with the RecoverEsupport intervention at the pre-specified ‘minimum’ dose and were included in the per-protocol analysis. The median length of stay was 5 days in the intervention group and 6 days in the control group. Participants in the intervention group stayed for 6% fewer days compared to participants in the control group (Table 2) (RR=0.94; 95% CrI 0.73-1.22; Pr(Superiority)=0.677), however the strength of the evidence was weak.

#### Secondary Outcomes

38 of 57 (67%) participants completed the in-hospital survey, and with completion rates slightly lower in the intervention group than the control group (57% vs 76% respectively). Those who did and did not complete the in-hospital survey were similar with respect to gender, stoma and open/laparoscopic surgery, however, completers were younger (mean age: completer=62, non-completer=70) and more likely to come from the private hospital (58%) than the public hospital (42%).

### ERAS BEHAVIOURS AND KNOWLEDGE (Day 1 post-surgery)

There were three behaviours for which the probability of intervention superiority was greater than 90% representing moderate or stronger evidence of an intervention effect. Relative to controls, intervention patients spent 2.38 times as long out of bed (RR=2.38) corresponding to a median of 195 and 60 minutes out of bed for the intervention and control groups respectively, with strong evidence of superiority (Pr(Superiority)=1.000). Similarly, relative to controls, intervention patients spent 1.28 times more moving (RR=1.28; Pr(Superiority)=0.904), corresponding to a median of 23 and 20 minutes respectively. Furthermore, intervention patients had higher odds of drinking at least 4 cups of fluid (OR=5.67; Pr(Superiority)=0.987), corresponding to 75% of intervention patients, compared to 45% of control patients meeting this threshold. With the exception of the proportion of participants with a PCA Device, all other ERAS behavioural and knowledge outcomes favoured the intervention group relative to controls (Table 2), albeit with weaker evidence of an intervention effect (Pr(Superiority) <0.900).

#### Quality of Recovery

Patient-rated Quality of Recovery scores were on average 11.64 points higher in the intervention group relative to controls (Pr(Superiority)=0.937), representing moderate evidence of intervention superiority.

#### Engagement

On average, intervention patients reported that they logged onto the website 10 times and spent an average total of 163 minutes on the website and 93% (13/14) completed at least one diary while in hospital, with 73% of the requested daily diaries completed while in hospital.

## DISCUSSION

This RCT evaluated the effectiveness of the RecoverEsupport DHI in improving the in-hospital recovery outcomes following colorectal cancer surgery. While no effect on median length of hospital stay was observed, there was moderate to strong evidence that the RecoverEsupport intervention was superior to usual care in terms of patient-reported quality of recovery, early mobilisation (both time spent out of bed and time spent moving) and early fluid intake (proportion consuming ≥4 cups). These findings suggest that digitally delivered behavioural support may improve certain aspects of post-operative recovery.

The trial was underpowered for detecting the prespecified LOS effect, and therefore estimates should be interpreted as preliminary. The absence of a decrease in median length of stay may also reflect that LOS is a distal outcome influenced by many patient-, clinical-, and system-level factors, and therefore may be less responsive to changes in the specific recovery behaviours targeted by the intervention. Results are broadly comparable to previous trials of DHIs to improve adherence to ERAS behaviours among CR surgical patients. Previous RCTs^13,11^ and non-randomised trials^12^ found no between-group difference in median length of stay. However, contrary to current results, the non-randomised by Schlund et al study found a decrease in mean LOS of ∼2 days.^12^ Given LOS data following surgery is typically skewed (including in the current study which included an extreme value of 62 days in the intervention group), it is important to distinguish between effects on the mean and median length of stay. A reduction in mean LOS can result from decreases in a small number of prolonged admissions, whereas the median better reflects the typical patient experience and may be harder to shift.

The between-group difference in time spent out of bed was substantial, with intervention participants spending 195 minutes out of bed, 2.38 times the time spent out of bed by controls, with strong evidence of intervention superiority (Pr(superiority)=1.00). Intervention participants also spent more time moving than control participants (1.28 times the minutes spent moving), although the difference was much smaller. It is difficult to contextualise the current results given previous studies have defined mobilisation differently (e.g. step counts^10^ vs proportion adherent^11,13^) and measured the behaviour over different time periods, and found conflicting results.^11,13^ Interestingly Van der Storm et al, also found that significant increases in mobilisation did not lead to any decrease in median length of hospital stay for the intervention group.^11^

Increases in mobilisation may also contribute to other physical, functional and psychological aspects of recovery. For example, while causality cannot be established, it is plausible that the improvements in mobilisation contributed to the observed improvements in quality of recovery scores. Intervention participants reported QoR-15 scores on average 11.64 points higher than controls. This exceeds the established clinically meaningful difference on this scale (∼ 8 points).^27^ Importantly, because the QoR-15 measures multiple domains (physical comfort, emotional state, physical independence, and psychological support) this improvement may represent broad improvement, rather than improvement in a single area.

Study strengths include its rigorous RCT design, objective measurement of the primary outcome using medical record data, and the use of an intervention grounded in behavioural science and informed by clinical stakeholders and patient input, with evidence of meaningful engagement. However, several limitations should be acknowledged. Recruitment fell short of the pre-specified sample size due to numerous reasons including staff changes and theatre closures. Although a Bayesian approach is not as dependent on sample size as a frequentist approach, the smaller sample size resulted in less participant variability and made it less likely that the full benefit of randomisation was realised, with intervention patients tending to be less well (i.e. a higher proportion classified as ASA III) relative to controls. Furthermore, some behavioural outcomes were assessed using self-reported items created specifically for this study with unknown psychometric properties, and the full range of analytic data from the website was not accessible.

In addition to a larger-scale multi-centre trial to confirm whether these findings generalise across different contexts, future research should also investigate the potential to embed DHIs such as RecoverEsupport within electronic medical record systems or existing digital hospital infrastructure, which would facilitate scalable integration into routine perioperative care pathways.^28^

In conclusion, although the RecoverEsupport intervention did not reduce length of hospital stay, it improved adherence to several patient-led ERAS behaviours (time spent out of bed, time spent moving, and fluid intake) and increased quality of recovery. These findings suggest that behaviourally-informed DHIs can improve adherence to recovery behaviours and may improve quality of recovery, highlighting the potential of digitally-delivered behavioural interventions as an adjunct to standard surgical care.

## Supporting information

Supp file 1 - CONSORT checklist

Supp file 2 - BCTs

Supp file 3 - Survey items

Supp file 4 - descriptive statistics

## Data Availability

All data produced in the present study are available upon reasonable request to the authors

## Author contributions

RW, KF, SS, SR and SAJ developed the research protocol. RW, KF, SAJ, PVDS, SS and SR developed and refined the intervention. SAJ and JC recruited patients, delivered the intervention and extracted the data. RW and PVDS managed the trial, and APP, PVDS and RW managed the data. EN and LL conducted the statistical analysis. All authors contributed to and approved the final version of this manuscript.

## Acknowledgements

The authors wish to thank Dr Alison Zucca for her role as a senior research assistant on the trial, Patrick Skippen for his role in developing the REDCap database, Sarah Leask for her advice about statistical analysis, and Cecilia King for her role in patient recruitment and intervention delivery. Furthermore, we grateful acknowledge the preliminary research, advice, support and guidance from Elise Mansfield, Professor Robert Sanson-Fisher, Professor Jon Gani, Professor Mariko Carey, Professor Andrew, Searles, Associate Professor Peter Pockney, Professor Christopher Oldmeadow, Associate Professor Ross Kerridge for their support and advice. We also acknowledge the support provided by the Equity in Health and Wellbeing Research Program and the HMRI Population Health Research Program.

## REFERENCES

1 Bray F, Laversanne M, Sung H, et al. Global cancer statistics 2022: GLOBOCAN estimates of incidence and mortality worldwide for 36 cancers in 185 countries. CA Cancer J Clin 2024; 74: 229–63.

2 Laparoscopic surgery for colorectal cancer. NICE (National Institute for Health and Care Excellence), 2006 https://www.nice.org.uk/guidance/ta105/resources/laparoscopic-surgery-for-colorectal-cancer-pdf-82598014092229 (accessed Aug 31, 2026).

3 Guidelines - ERAS® Society. https://erassociety.org/guidelines/ (accessed July 16, 2026).

4 Greco M, Capretti G, Beretta L, Gemma M, Pecorelli N, Braga M. Enhanced Recovery Program in Colorectal Surgery: A Meta-analysis of Randomized Controlled Trials. World J Surg 2014; 38: 1531–41.

5 Sauro KM, Smith C, Ibadin S, et al. Enhanced Recovery After Surgery Guidelines and Hospital Length of Stay, Readmission, Complications, and Mortality: A Meta-Analysis of Randomized Clinical Trials. JAMA Netw Open 2024; 7: e2417310.

6 Gustafsson UO, Scott MJ, Hubner M, et al. Guidelines for Perioperative Care in Elective Colorectal Surgery: Enhanced Recovery After Surgery (ERAS®) Society Recommendations: 2018. World J Surg 2019; 43: 659–95.

7 van Zelm R, Coeckelberghs E, Sermeus W, et al. Variation in care for surgical patients with colorectal cancer: protocol adherence in 12 European hospitals. Int J Colorectal Dis 2017; 32: 1471–8.

8 Low CA, Danko M, Durica KC, et al. A Real-Time Mobile Intervention to Reduce Sedentary Behavior Before and After Cancer Surgery: Usability and Feasibility Study. JMIR Perioper Med 2020; 3: e17292.

9 Talen AD, Leenen JPL, Van Der Sluis G, Oldenhuis HKE, Klaase JM, Patijn GA. Feasibility of a Comprehensive eCoach to Support Patients Undergoing Colorectal Surgery: Longitudinal Observational Study. JMIR Perioper Med 2025; 8: e67425.

10 Jones ASK, Kleinstäuber M, Akroyd A, et al. Using animated visualization to improve postoperative mobilization: A randomized controlled trial. Health Psychol 2019; 38: 748–58.

11 Van Der Storm SL, Jansen M, Mulder MD, et al. Improving Enhanced Recovery after Surgery (ERAS): The Effect of a Patient-Centred Mobile Application and an Activity Tracker on Patient Engagement in Colorectal Surgery. Surg Innov 2025; 32: 5–15.

12 Schlund D, Poirier J, Bhama AR, et al. Value of an interactive phone application in an established enhanced recovery program. Int J Colorectal Dis 2020; 35: 1045–8.

13 Mata J, Pecorelli N, Kaneva P, et al. A mobile device application (app) to improve adherence to an enhanced recovery program for colorectal surgery: a randomized controlled trial. Surg Endosc 2020; 34: 742–51.

14 Wyse R, Smith S, Zucca A, et al. Effectiveness and cost-effectiveness of a digital health intervention to support patients with colorectal cancer prepare for and recover from surgery: study protocol of the RecoverEsupport randomised controlled trial. BMJ Open 2023; 13: e067150.

15 Michie S, Richardson M, Johnston M, et al. The behavior change technique taxonomy (v1) of 93 hierarchically clustered techniques: building an international consensus for the reporting of behavior change interventions. Ann Behav Med 2013; 46: 81–95.

16 Myles PS, Shulman MA, Reilly J, Kasza J, Romero L. Measurement of quality of recovery after surgery using the 15-item quality of recovery scale: a systematic review and meta-analysis. Br J Anaesth 2022; 128: 1029–39.

17 Schlund D, Poirier J, Bhama AR, et al. Value of an interactive phone application in an established enhanced recovery program. Int J Colorectal Dis 2020; 35: 1045–8.

18 Kass RE, Raftery AE. Bayes Factors. J Am Stat Assoc 1995; 90: 773–95.

19 van Buuren S, Groothuis-Oudshoorn K. mice: Multivariate Imputation by Chained Equations in R. J Stat Softw 2011; 45: 1–67.

20 Bürkner P-C. Advanced Bayesian Multilevel Modeling with the R Package brms. R J 2018; 10: 395–411.

21 Bürkner P-C. Bayesian Item Response Modeling in R with brms and Stan. J Stat Softw 2021; 100: 1–54.

22 Bürkner P-C. brms: An R Package for Bayesian Multilevel Models Using Stan. J Stat Softw 2017; 80: 1–28.

23 Stan Development Team. Stan: Software for Bayesian Data Analysis. Stan. https://mc-stan.org/ (accessed March 3, 2026).

24 Barret L, Liaigre L, Hlavaty A, et al. Priors and decision thresholds in phase 2 and phase 3 randomized controlled trials evaluating drug efficacy using Bayesian methods: a systematic review. J Clin Epidemiol 2025; 179: 111651.

25 Huang P, Zhu K, Wu T, et al. Ultrasound-guided intermediate cervical plexus block versus local infiltration anesthesia for thermal ablation of benign thyroid nodules: a non-inferior, double-blinded, randomized controlled trial. BMC Anesthesiol 2025; 25: 586.

26 Efficient LOO-CV and WAIC for Bayesian models — loo-package. https://mc-stan.org/loo/reference/loo-package.html (accessed Aug 31, 2026).

27 Myles PS, Myles DB. An Updated Minimal Clinically Important Difference for the QoR-15 Scale. Anesthesiology 2021; 135: 934–5.

28 Greenhalgh T, Wherton J, Papoutsi C, et al. Beyond Adoption: A New Framework for Theorizing and Evaluating Nonadoption, Abandonment, and Challenges to the Scale-Up, Spread, and Sustainability of Health and Care Technologies. J Med Internet Res 2017; 19: e367.

