## Supplementary material for "The effectiveness of the ‘RecoverEsupport’ digital health intervention in enhancing recovery from colorectal cancer surgery: RCT outcomes from the hospital admission": Supp file 2 - BCTs

**Supplementary File 2:**
Behaviour change strategies used in the RecoverEsupport intervention based on the COM-B model(1) and the framework for digital behaviour change strategies by Wang et al.(2) Table adapted from the RecoverEsupport protocol(3)

| **Behaviour change strategy & description  (from Yang et al.)** | **How strategy is operationalised in ‘RecoverEsupport’** |
| --- | --- |
| **STRATEGIES TO SUPPORT INTERVENTION ENGAGEMENT** | |
| **Prompts / Cues:** Introduce or define environmental or social stimulus with the purpose of prompting or cueing the behaviour. The prompt or cue would normally occur at the time or place of performance. | - **Surgeon’s ‘Prescription Letter’** - Intervention participants received a letter from the surgical services team prescribing the RecoverEsupport program and to prompt patients to access the website and increase engagement. - **Patient prompts** - Patients received a series of personalised automated reminders to prompt them to access the website (pre-and post-surgery) and complete daily diaries (in hospital). - **Clinician alerts** - When patient responses in the ‘Daily Diary’ indicated non-adherence to the ERAS recommendations, or when patients flag they were distressed, the website sent an email alert to the Colorectal Cancer Liaison Nurse to follow-up with the patient within the day. - **Patient Questions** - ‘My Questions’ - At the end of each website module, patients were asked if they had questions for their clinical team. Patients could record and save their questions centrally within the website and email a copy to themself and/or the Colorectal Cancer Liaison Nurse. Patients were encouraged to print and/or bring their list of questions to medical appointments. - **Post-discharge care** - At discharge, the patient’s GP was faxed a list of common side effects/complications following colorectal surgery and how to manage them. The patient also received a reminder to make a follow-up appointment with their GP 2 weeks after discharge, and were invited to record that they had done this. - **Provision of Device** - To increase website engagement during the inpatient stay, intervention patients without access to a smartphone or tablet or laptop were provided one during their hospital stay (public hospital patients only). |
| **Social support:** Advise on, arrange or provide social support (e.g. from friends, relatives, colleagues or staff) | - **Support Person involvement** - The support person could access the website independently of, or together with the patient. - **GP involvement -** On discharge, a standard information sheet detailing the general care needs for CRC patients was faxed to the patient’s GP. |
| **STRATEGIES TO MOTIVATE BEHAVIOUR CHANGE** | |
| **Credible source:**  Present verbal or visual communication from a credible source in favour of or against the behaviour | Information was communicated via videos presented by the clinical team who were providing care to the patient, including a surgeon, anaesthetist, and stoma nurse. There were also a series of videos of CRC patients talking first-hand about their experiences. |
| **Framing/reframing:**  Suggest the deliberate adoption of a new perspective on behaviour (e.g. its purpose) in order to change cognitions or emotions about performing the behaviour | The website encouraged patients to see themselves as active participants in their own recovery, to empower them to take control of their recovery. Patients were asked to reflect on their motivation for accessing the intervention and optimising their recovery. |
| **Information about health consequences:** Provide information (e.g. written, verbal, visual) about health consequences of performing the behaviour | Text and videos were included within the website outlining the rationale for undertaking the behaviours specified in the ERAS recommendations, and the benefits of adherence and the consequences of non-adherence were explained, e.g. If you remain on opioids, your bowel will take longer to start working again. |
| **STRATEGIES TO ENACT BEHAVIOUR CHANGE** | |
| **Self-monitoring of behaviour:** Establish a method for the person to monitor and record their behaviour/s as part of a behaviour change strategy | **Daily diary:** Each day in the hospital post-surgery, the patient was prompted (by SMS/email) to use the website to monitor and record their behaviours that support recovery (e.g. moving, eating, drinking, breathing exercises, and minimising opioids). |
| **Feedback on behaviour:** Monitor and provide informative or evaluative feedback on the performance of the behaviour | **Daily diary:**   - The website provided automated and tailored feedback on patient behaviour (e.g. mobilisation) based on patient self-monitoring data (see above). - Non-adherence to ERAS recommendations triggered an alert to the Colorectal Cancer Liaison Nurse so that they could follow-up with the patient (i.e. if they were not getting up and moving) and attempt to address barriers   **Interactive quizzes:** Brief self-assessment tools (with real-time feedback) were included in each module to ensure patients understand:   - What the key patient-led ERAS recommendations were (e.g. opioid minimisation post-surgery) - Why they were important to their recovery (e.g. quickens return to normal bowel function and defecation) and   What they could do (e.g. take all non-opioid analgesia according to the prescribed schedule). |
| **Instruction and demonstration of how to perform the behaviour:**  Advise how to perform the behaviour (including the provision of an observable sample of the performance of the behaviour) | Patients were provided with text, videos and diagrams explaining step-by-step how to undertake specific behaviours.  Specifically, diagrams and videos were included on the website to demonstrate   - the target behaviours, e.g. breathing exercises. - What to expect at each stage of the patient journey, e.g. what patients see, hear and feel as they are taken into the operating theatre. |

**REFERENCES**

1.	Michie S, Richardson M, Johnston M, Abraham C, Francis J, Hardeman W, et al. The behavior change technique taxonomy (v1) of 93 hierarchically clustered techniques: building an international consensus for the reporting of behavior change interventions. Ann Behav Med. 2013 Aug;46(1):81–95. doi:10.1007/s12160-013-9486-6 PubMed PMID: 23512568.

2.	Wang Y, Fadhil A, Lange JP, Reiterer H. Integrating Taxonomies Into Theory-Based Digital Health Interventions for Behavior Change: A Holistic Framework. JMIR Research Protocols. 2019 Jan 15;8(1):e8055. doi:10.2196/resprot.8055

3.	Wyse R, Smith S, Zucca A, Fakes K, Mansfield E, Johnston SA, et al. Effectiveness and cost-effectiveness of a digital health intervention to support patients with colorectal cancer prepare for and recover from surgery: study protocol of the RecoverEsupport randomised controlled trial. BMJ Open. 2023;13(3):e067150. doi:10.1136/bmjopen-2022-067150 PubMed PMID: 36878662; PubMed Central PMCID: PMC9990701.
