## Supplementary material for "The effectiveness of the ‘RecoverEsupport’ digital health intervention in enhancing recovery from colorectal cancer surgery: RCT outcomes from the hospital admission": Supp file 3 - Survey items

**Supplementary File 3:**

In-hospital online survey assessing ERAS behaviours and knowledge:

**** Please note, “the day after surgery” refers to the first full day that you spent on the ward after your surgery. ****

**Behaviours**

*The day after your surgery, how much time did you spend out of bed?*

* Include any time you were sitting in the chair, moving around inside or outside of the ward, dressing, showering etc. (Hours and/or minutes)

*The day after your surgery, how many hours did you spend walking or moving around?*

* Include time spent moving around inside or outside of the ward, dressing, showering (Hours and/or minutes)

*The day after your surgery, how many cups of liquid did you drink?*

*Include all fluids – water, cordial, juice, tea, coffee, Ensure® / Resource® supplements etc. A standard 600ml water bottle holds approximately 2.5 cups*

*The day after your surgery, how many meals did you eat at least some of? (i.e. Breakfast, lunch, dinner)*

3 or more / 2 / 1 / 0

*The day after your surgery, how many snacks did you attempt to eat at least some of? (i.e. Morning tea, afternoon tea, supper)*

3 or more / 2 / 1 / 0

*The day after your surgery, during your waking hours, how many times did you deliberately deep breath, cough or huff and puff each day?*

Every hour / Every 2-3 hours / Every 4-5 hours / Once or twice / Not at all

*The day after your surgery, did you have a patient-controlled device to administer pain medication?*

Yes / No / Don’t Know

**Knowledge:**

**To what extent do you agree or disagree with the following statements:**

*I feel as though I am able to do things to enhance or improve my recovery*

Strongly agree / Agree / Neither Agree nor Disagree / Disagree / Strongly Disagree

I understand the things I need to do to enhance/improve my recovery

Strongly agree / Agree / Neither Agree nor Disagree / Disagree / Strongly Disagree
