## Supplementary material for "The effectiveness of the ‘RecoverEsupport’ digital health intervention in enhancing recovery from colorectal cancer surgery: RCT outcomes from the hospital admission": Supp file 4 - descriptive statistics

**Supplementary File 4:**

Median intervention vs control hospital length of stay (days) by gender, age group and hospital type

|  | **Intervention (RecoverEsupport)**  **N=28** | **Control (Usual Care)**  **N=29** |
| --- | --- | --- |
| **GENDER** (median (N, IQR)) |  |  |
| Female | 7 (*16*, 4, 8.5) | 6 (*15*, 5, 12.5) |
| Male | 5 (*12*, 4, 8.5) | 5 (*14*, 4, 6.75) |
| **AGE GROUP** (median (N, IQR)) |  |  |
| ≤ 67 years | 7 (*12*, 4, 10.25) | 6 (*17*, 5, 8) |
| > 67 years | 5 (*16*, 4, 7) | 5 (*12*, 4, 8) |
| **HOSPITAL TYPE** (median (N, IQR)) |  |  |
| Public | 7 (*13*, 5, 11) | 6 (*14*, 5, 11.75) |
| Private | 4 (*15*, 4, 7) | 6 (*15*, 4, 6.5) |
